# Harnessing AI and social media to understand real-world patient experiences in systemic lupus erythematosus

**DOI:** 10.64898/2026.02.20.26346724

**Authors:** Siqi Yang, Carina Hawryluk, James Liu, Niki Eckert, Jessica Otoo, Ernest R. Vina, Lixia Yao

## Abstract

**Objective:** To apply large language models (LLMs) to Reddit posts referencing systemic lupus erythematosus (SLE) to identify patient-expressed unmet medical needs, symptom experiences, and healthcare challenges, demonstrating how AI-enabled social media listening complements traditional patient-experience research.

**Methods:** We extracted 4,633 posts from ten SLE-related or health-focused Reddit communities using the public Reddit API (October–November 2025). After removing duplicates, promotional content, and posts with insufficient information, 2,603 posts remained. A thematic codebook was developed through manual review of 300 posts and iteratively refined. Two LLMs (Gemini 3.0 and GPT-5.2) were evaluated for automated thematic labeling using percent agreement, Cohen’s κ, and a human-annotated reference set (n=100). The best-performing model was used to quantify theme prevalence, followed by qualitative review of representative narratives.

**Results:** GPT-5.2 demonstrated higher performance (F1=0.844) than Gemini 3.0 (F1=0.811), with substantial inter-model agreement across main themes (mean κ=0.71). Posts reflected multidimensional experiences. The most frequent subtheme was *Advice Seeking* (84.1%), followed by *Emotional Coping* (55.6%). Common symptom-related themes included *Pain* (37.2%), *Other Symptom Presentations* (37.6%), *Fatigue* (24.7%), and *Acute or Worsening Flares* (30.2%). Diagnostic uncertainty was prominent, including confusion about laboratory results (24.0%) and emotional impact of uncertainty (33.0%). Qualitative review highlighted emotional distress, reliance on peer communities for interpretation of symptoms and labs, and difficulty managing complex treatment regimens.

**Conclusion:** LLM-enabled social media listening offers a scalable method for synthesizing large volumes of unstructured patient narratives, providing timely insights into lived experiences and unmet needs among individuals discussing lupus online. Findings align with established qualitative literature while highlighting persistent gaps in patient education, communication, and care coordination. This analytical framework can be applied across disease areas to support patient-centered care, measurement development, and evidence generation relevant to therapeutic and health-services research.

**What is already known on this topic:**

- People living with systemic lupus erythematosus (SLE) experience substantial unmet needs related to diagnostic uncertainty, symptom burden, emotional distress, medication challenges, and healthcare system barriers.
- Traditional qualitative methods (e.g., interviews, focus groups, surveys) capture valuable patient perspectives but are limited by small sample sizes, recall bias, and restricted question frameworks.
- Social media listening has emerged as a promising way to collect real-time patient insights, and recent regulatory guidance acknowledges its value as patient experience data. However, systematic, scalable analysis of large patient-generated datasets has historically been constrained by analytic burden and variability.

**What this study adds:**

- This study is among the first to apply **state-of-the-art large language models (LLMs)** to a large corpus of SLE-related social media posts, enabling scalable thematic analysis of thousands of patient narratives.
- It provides **a validated methodological framework** for using dual-LLM agreement, human-annotated references, and performance benchmarking (precision, recall, F1) to ensure reliability in automated thematic labeling.
- Findings reveal a **multidimensional patient burden** consistent with prior studies while uncovering persistent gaps in patient education, confusion around laboratory testing, care coordination challenges, and heavy reliance on peer communities for advice.
- The approach demonstrates that LLM-enabled social media listening can generate timely, granular, patient-prioritized insights at a scale unattainable by traditional methods.

**How this study might affect research, practice, or policy:**

- **Research:** Establishes a reproducible, scalable framework for integrating LLM-based thematic analysis into patient-focused evidence generation, accelerating insight extraction from large unstructured datasets across disease areas.
- **Clinical practice:** Highlights actionable gaps in patient education, communication, and care coordination, informing interventions to improve clinical encounters, shared decision-making, and symptom management support.
- **Policy and regulatory science:** Demonstrates how social media–derived patient experience data, when paired with rigorous quality controls, can complement formal qualitative studies and support patient-focused drug development, measurement development, and health-services planning.

## INTRODUCTION

Systemic Lupus Erythematosus (SLE) is a complex, chronic autoimmune disease characterized by immune-mediated inflammation affecting multiple organ systems.^1^ Globally, the incidence rate of SLE is estimated at 5.14 per 100,000 person-years ^2^, with approximately 90% of cases occurring in females.^1^ The disease exhibits profound heterogeneity in clinical presentation, disease trajectory, and immunological features, which complicates diagnosis, delays appropriate treatment, and contributes to persistent disparities in care.^3,4^ Together, these challenges impose a substantial and multifaceted burden on individuals living with SLE.

Despite substantial treatment advances over the past several decades ^5,6^, critical gaps remain in our understanding of disease burden and treatment from the patient perspective. Physician-assessed disease activity and biomarker-based evaluations often fail to align with patient-reported health status, symptom severity, and quality of life.^7^ This discordance reflects a fundamental dilemma of current clinical assessment frameworks: they tend to prioritize objectively measurable disease activity over the lived experience of illness while the biological mechanisms of SLE are only partly understood and biomarkers may not consistently capture symptom burden. Consequently, important dimensions of unmet medical needs, such as symptom fluctuation, treatment trade-offs, psychosocial burden, and disruptions to daily functioning remain underrecognized or insufficiently characterized. Addressing this gap is essential for advancing patient-centered care and informing therapeutic development that meaningfully reflects patient priorities.^8–10^

Traditionally, patient experience data has been collected through structured methods such as interviews, focus groups, and surveys.^11,12,13^ While these approaches provide valuable insights, they are inherently limited by small sample sizes, recall bias, and the constraints of predefined questions.^11,12,13^ These limitations are particularly pronounced in heterogeneous conditions like SLE, where recruitment can be challenging and patient experiences vary widely across different disease phenotypes and life stages. Thus, traditional methods may only capture a partial and static view of patient burden, rather than the dynamic and evolving nature of living with SLE.

In today’s digital age, social media platforms have emerged as invaluable repositories of real-time patient insights and experiences.^14,15^ They offer unprecedented access to a wealth of unfiltered, candid narratives from individuals living with chronic diseases like SLE. Through social media listening, researchers and healthcare providers gain unique opportunities to identify and address previously unrecognized patient needs and concerns. Recognizing the potential of such data, regulatory bodies like the U.S. Food and Drug Administration (FDA) have officially acknowledged social media as a legitimate source of qualitative and quantitative patient experience data, providing guidelines on its ethical use starting in 2022.^16^

The integration of advanced natural language processing techniques, including large language models (LLMs), has revolutionized how we interpret and utilize these vast datasets.^17,18^ Multiple studies have applied LLMs to social media data across various diseases, including evaluation of glaucoma treatment^19^, analyses of unmet needs and provider burnout in thyroid disease^20^, and assessments of public perceptions of obesity treatment^21^.

In this study, we apply LLM-enabled social media listening to systematically analyze Reddit posts from lupus-related communities, aiming to generate a comprehensive, patient-prioritized map of unmet medical needs and lived experiences in SLE. This approach provides a scalable and timely complement to patient-focused evidence generation, with direct relevance to care delivery, patient education, and shared decision-making. In addition, it establishes a reproducible analytical framework for leveraging patient-generated data to inform measurement development and therapeutic strategies.

## METHODS

### Data collection and preprocessing

This study used the free, publicly available Reddit Application Programming Interface (API) to retrieve patient-generated content related to SLE from ten subreddits (Figure 1). Subreddits are topic-specific forums on Reddit in which users create posts and engage in threaded discussions. Two complementary data retrieval strategies were employed. First, we collected posts published from eight lupus-specific subreddits: r/lupus, r/lupussupport, r/lupusRantAndRelief, r/lupusResearch, r/lupusmicrobiome, r/ItsNeverlupus, r/menwithlupus, and r/Livingwithlupus. Second, we conducted keyword-based searches (“lupus” OR “systemic lupus erythematosus”) within two broader health-focused subreddits (r/autoimmune and r/chronicillness). Data extraction was conducted between October 14, 2025 and November 25, 2025, yielding a total of 4,633 posts.

**Figure 1.**
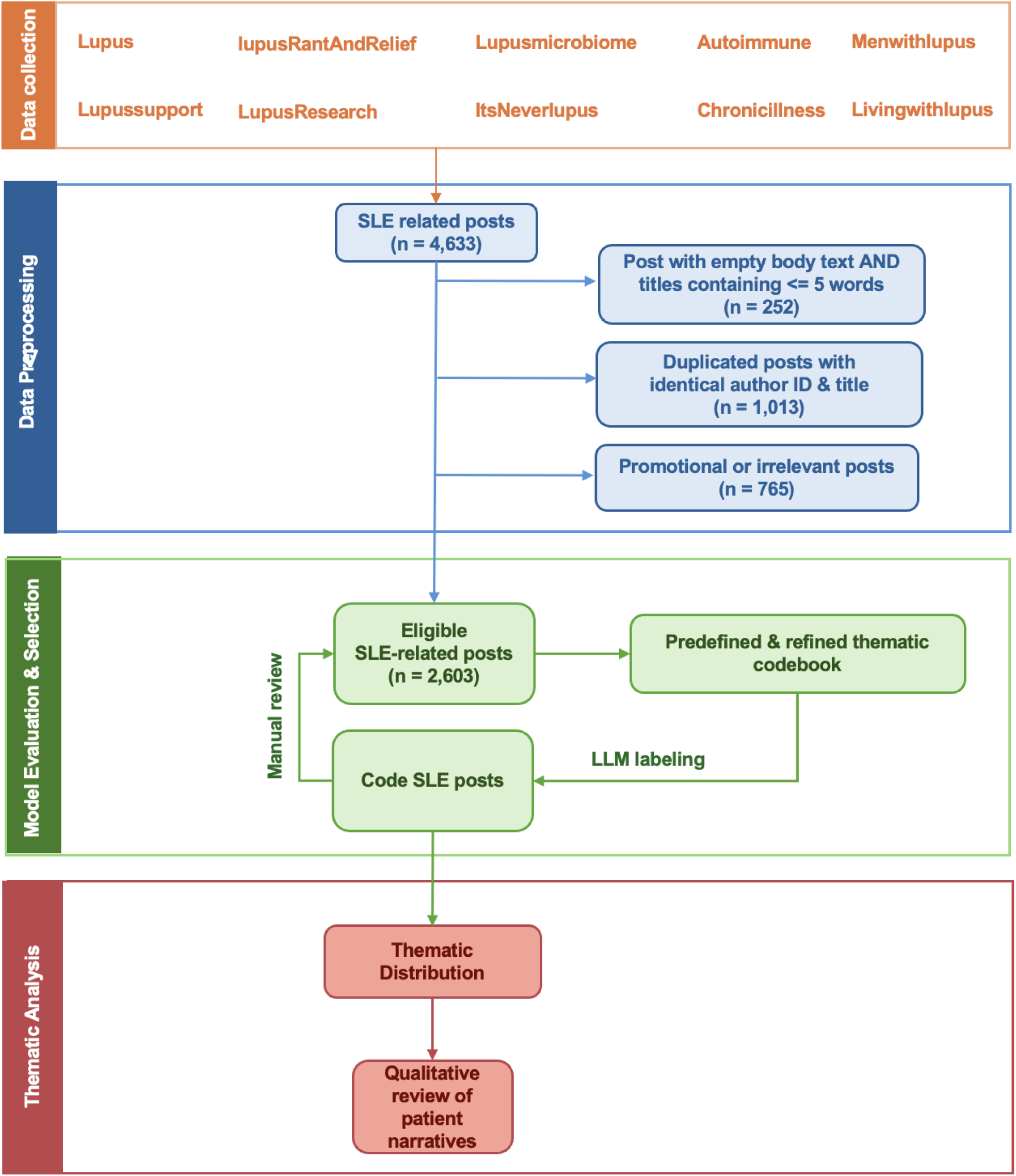
Workflow diagram for data collection, preprocessing, and LLMl1Jbased thematic analysis. **(A)** Main themes consistency distribution **(B)** Subthemes consistency distribution

To ensure data quality and relevance, we implemented a multi-step preprocessing pipeline. Posts that contained both empty body text and title of five or fewer words were excluded due to insufficient context to reliably reflect patient or caregiver experiences. Duplicate posts were identified and removed using an exact-match rule based on identical author identifiers and post titles. Promotional materials (e.g., advertisement or research recruitment posts) and other non-patient reported content were identified and excluded with assistance from two state-of-the-art LLMs (Google Gemini 3.0 and OpenAI GPT-5.2).

### Thematic codebook development

A thematic codebook was developed to systematically classify patient experiences and unmet medical needs. Three public health researchers independently reviewed and manually annotated 300 randomly selected posts to inductively generate an initial set of themes and subthemes. To ensure comprehensive coverage of the full dataset, an “Other” category was incorporated into the preliminary codebook, which was then used for an initial round of automated labeling across all posts. This step allowed for the identification of content not adequately captured by the initial thematic structure. If no “Other” category was identified in the entire dataset, we remove “Other” category from the final results. If no posts were assigned to the “Other” category across the full dataset, the category was excluded from the final codebook.

Following a review of a sampled model outputs, the research team iteratively refined the codebook to improve definitional clarity, enhance conceptual coverage, and ensure mutual exclusivity between subthemes. The finalized codebook comprised six main themes and 43 subthemes (Table 1) and was subsequently applied for definitive automated thematic labeling of the full dataset.

**Table 1.** Percent agreement and Cohen’s κ between Gemini 3.0 Pro and GPTl7l5.2.

| Themes | Percent Agreement | Cohen's $\kappa$ |
| --- | --- | --- |
| Diagnosis Journey & Uncertainty | 86.86 | 0.74 |
| Delayed, Difficult, or Incorrect Diagnosis | 93.89 | 0.79 |
| Emotional Impact of Uncertainty and Disease Progression | 81.18 | 0.56 |
| Testing & Results Confusion (ANA, dsDNA, complements) | 93.12 | 0.82 |
| Provider Trust/ Distrust | 91.20 | 0.64 |
| Other | 92.20 | 0.21 |
| Emotional & Social Support | 98.27 | 0.42 |
| Advice Seeking | 96.27 | 0.86 |
| Emotional Coping (e.g., venting, hope, resilience) | 86.71 | 0.73 |
| Identity and/or Future Uncertainty | 89.70 | 0.68 |
| Relationship Strain / Interpersonal Challenges | 96.70 | 0.84 |
| Bereavement / Loss | 99.15 | 0.73 |
| Healthcare System Barriers | 89.01 | 0.76 |
| Access To Care (availability, wait times, transport) | 96.12 | 0.80 |
| Patient Education Gaps & External Misinformation | 90.43 | 0.50 |
| Dismissal Or Minimization By Providers | 95.70 | 0.85 |
| Insurance Challenges (coverage, prior auth) | 99.35 | 0.91 |
| Financial Burden Beyond Insurance | 98.16 | 0.81 |
| Care Coordination Challenges (referrals, records) | 95.01 | 0.65 |
| Clinical Care Errors / Safety Events | 98.85 | 0.62 |
| Medication Experiences & Side Effects | 94.08 | 0.88 |
| Medication Effectiveness / Response | 92.24 | 0.80 |
| Medication Side Effects / Adverse Events | 95.93 | 0.87 |
| Infusion Experience (e.g., infusion centers, scheduling) | 98.27 | 0.74 |
| Treatment Recommendations | 90.32 | 0.66 |
| Supplements Or Alternative Therapies | 98.12 | 0.83 |
| Treatment Adherence Challenges (forgetting, intolerance) | 96.24 | 0.63 |
| Biologics vs Traditional Medications | 96.77 | 0.68 |
| Clinical Trials Or Research Participation | 99.69 | 0.84 |
| Other | 96.39 | 0.16 |
| Symptoms & Physical Experiences | 92.24 | 0.76 |
| Acute or Worsening Flares | 88.78 | 0.73 |
| Fatigue | 96.77 | 0.91 |
| Pain | 94.43 | 0.88 |
| Skin Manifestations (rashes, photosensitivity) | 94.85 | 0.86 |
| Cognitive Symptoms (brain fog, memory) | 98.77 | 0.91 |
| Other Symptom Presentation (e.g., renal, cardio) | 84.48 | 0.69 |
| Other | 94.08 | 0.18 |
| Lifestyle & Self-Management | 85.79 | 0.72 |
| Symptom Management Strategies | 84.67 | 0.57 |
| Diet and Nutrition Changes | 97.77 | 0.85 |
| Exercise Adaptations | 98.35 | 0.81 |
| Stress Management Strategies | 96.39 | 0.53 |
| Workplace Disclosure / Accommodations & Disability Benefits | 96.62 | 0.80 |
| Sun Exposure Management | 97.69 | 0.80 |
| Self-Tracking / Tools (journals, apps) | 98.62 | 0.74 |
| Activities Of Daily Living Adjustments | 87.98 | 0.65 |
| Body Image / Identity Adjustments | 94.12 | 0.60 |
| Reproductive / Fertility Treatment Considerations | 98.73 | 0.83 |

*GPT, Generative Pre-trained Transformer; $\kappa$ , Cohen's Kappa coefficient*

### LLM evaluation and model selection

Two state-of-the-art LLMs, Google Gemini 3.0 and OpenAI GPT-5.2, were used for automated thematic labeling via their respective APIs. Prompt engineering was performed to optimize instructions for consistent labeling of Reddit posts to the main themes and subthemes defined in the final thematic codebook. Both models were instructed to assign all applicable main theme and subtheme labels for each post, rather than selecting a single dominant theme, to reflect the multidimensional nature of patient narratives.

To assess the reliability of automated thematic labeling, we evaluated inter-model consistency by calculating percent agreement and Cohen’s kappa (κ)^22^ across all main themes and subthemes (Table 1). Agreement was defined as both models assigning the same binary label (Yes or No) for a given theme or subtheme within a post. Interpretation of κ followed established empirical guideline: ≤ 0, indicating no agreement; 0.01–0.20, slight; 0.21–0.40, fair; 0.41– 0.60, moderate; 0.61–0.80, substantial; and 0.81–1.00, almost perfect agreement.^23^ The distribution of agreement across posts for both main themes and subthemes agreement was further visualized to characterize consistency patterns between the two LLMs (Figure 2).

**Figure 2.**
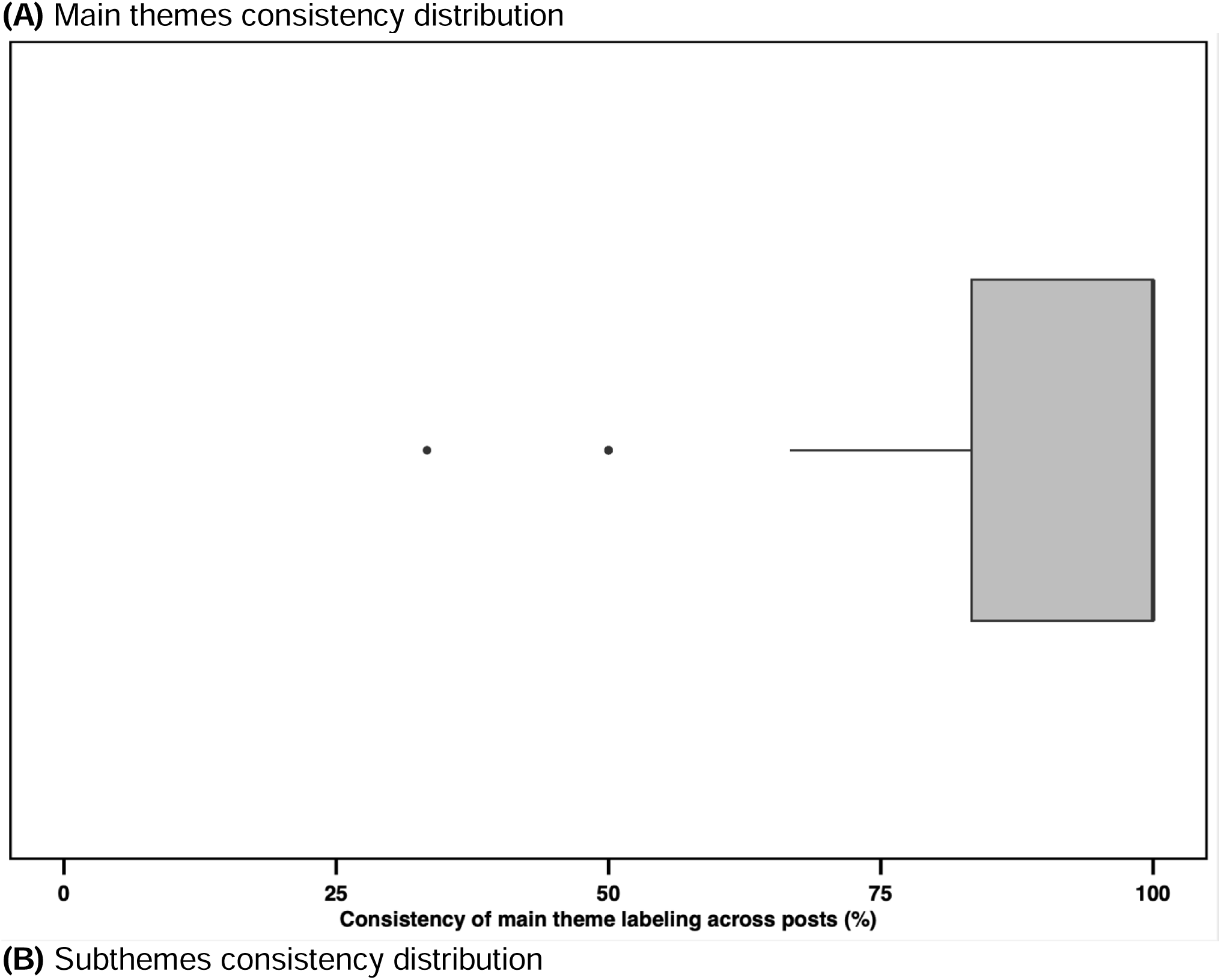
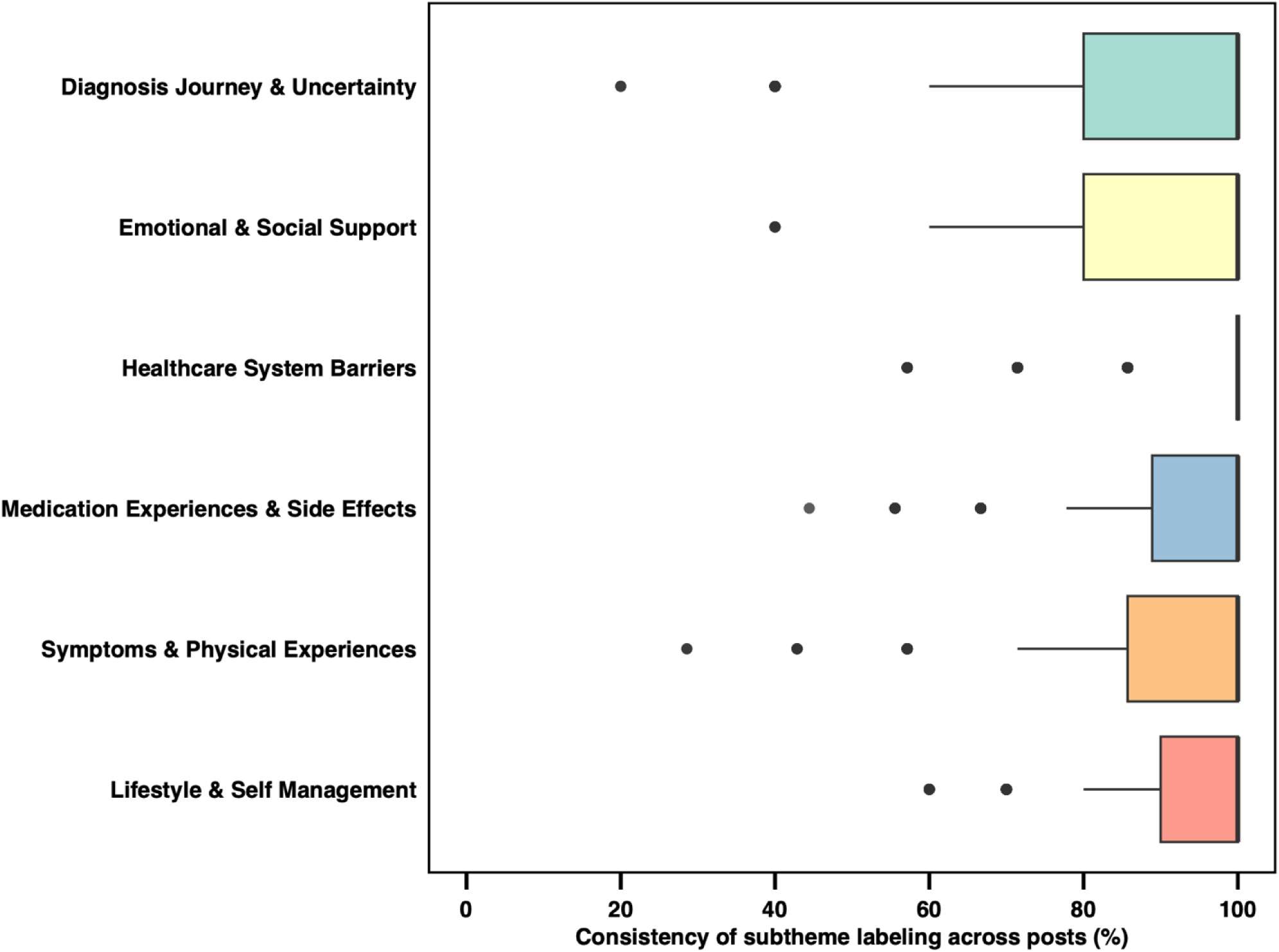
Thematic consistency of LLMs. **(A)** Boxplot showing the distribution of inter-model consistency for main theme labeling across posts. Consistency is defined as the percentage of posts assigned the same main theme label by Gemini 3.0 and GPT-5.2. The box denotes the interquartile range (25th–75th percentiles), and the median is shown as a line (overlapping the upper quartile in this panel). Outliers are shown as individual points. **(B)** Boxplots showing the distribution of inter-model consistency for subtheme labeling across posts, stratified by main theme. Consistency is defined as the percentage of posts assigned the same subtheme label by Gemini 3.0 and GPT-5.2. Boxes denote the interquartile range (25th–75th percentiles), and the median line overlaps the upper quartile in all panels. For Healthcare System Barrier, 25th is also overlapped with median and 75th percentile. Outliers are shown as individual points.

Additionally, three public health researchers independently annotated 100 randomly selected posts to establish a human-annotated reference dataset for performance evaluation. Discrepancies were resolved using a majority-vote consensus approach to determine the final labels for each main theme and subtheme. Using this consensus reference, model performance was evaluated using precision, recall, and F1-score, and the best-performing LLM was subsequently selected for downstream thematic analysis.

### Thematic analysis and reporting

Using the best-performing LLM, we quantified the prevalence of each main theme and subtheme by calculating counts and percentages across the 2,603 patient posts (Table 2). To support interpretation of the quantitative findings, we conducted a qualitative review of representative posts within each main theme to identify salient patient experiences and unmet medical needs. Selected examples were synthesized into key phrases and illustrative summaries to contextualize the thematic patterns observed and are presented in Figure 3.

**Figure 3.**
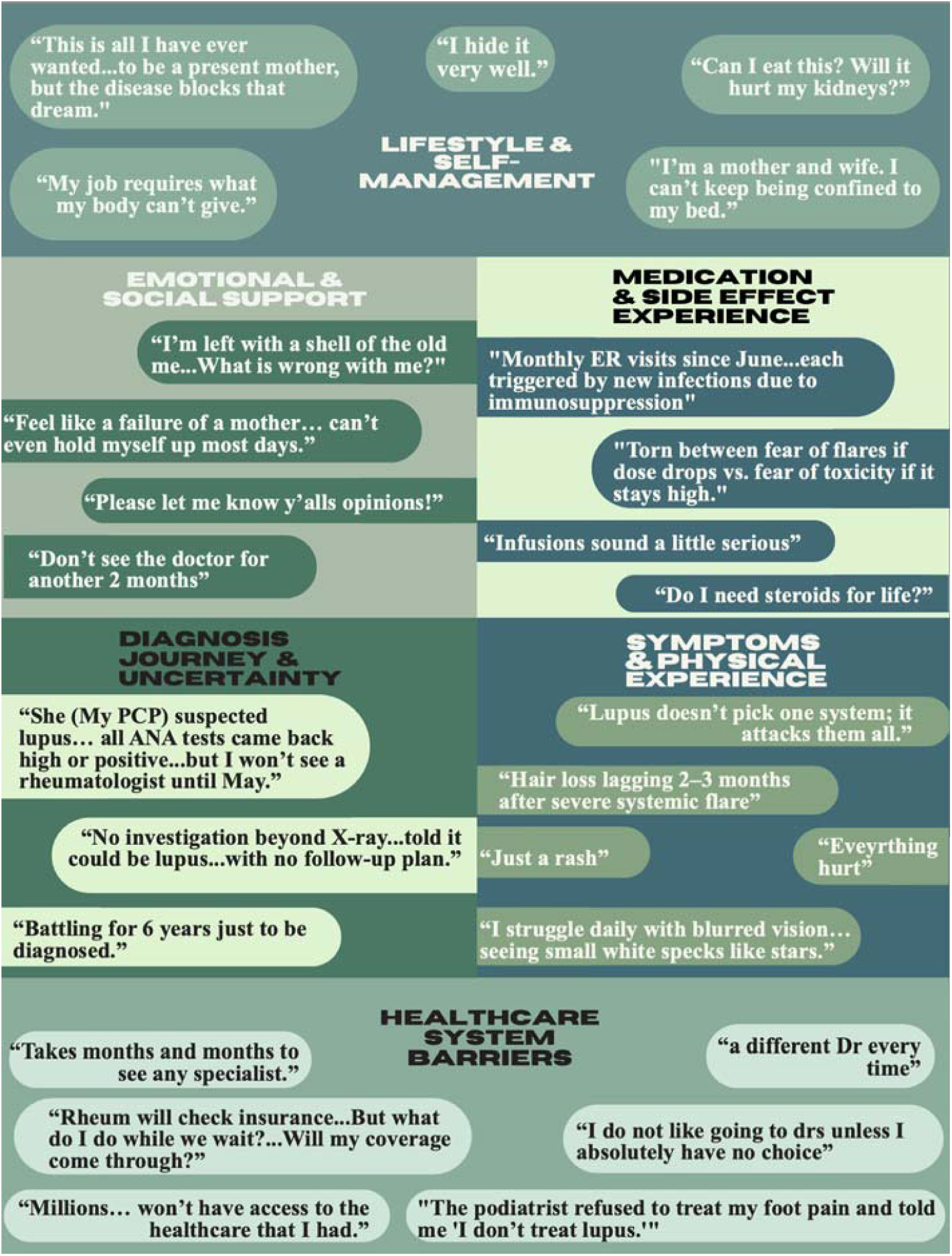
Unmet Medical Needs and Patient Experiences Illustrated by Selected Patient Narratives

**Table 2.** Distribution of main themes and subthemes identified by the bestl7lperforming model (OpenAI GPT 5.2)

| Main themes | Subthemes | OpenAI GPT 5.2 |
| --- | --- | --- |
| Diagnosis Journey & Uncertainty | Delayed, Difficult, or Incorrect Diagnosis | 432 (16.6%) |
|  | Emotional Impact of Uncertainty and Disease Progression | 860 (33.0%) |
|  | Testing & Results Confusion (ANA, dsDNA, complements) | 624 (24.0%) |
|  | Provider Trust/ Distrust | 426 (16.4%) |
|  | Other | 82 (3.2%) |
| Emotional & Social Support | Advice Seeking | 2190 (84.1%) |
|  | Emotional Coping (e.g., venting, hope, resilience) | 1448 (55.6%) |
|  | Identity and/or Future Uncertainty | 572 (22.0%) |
|  | Relationship Strain / Interpersonal Challenges | 278 (10.7%) |
|  | Bereavement / Loss | 47 (1.8%) |
| Healthcare System Barriers | Access To Care (availability, wait times, transport) | 290 (11.1%) |
|  | Patient Education Gaps & External Misinformation | 332 (12.8%) |
|  | Dismissal Or Minimization By Providers | 424 (16.3%) |
|  | Insurance Challenges (coverage, prior auth) | 104 (4.0%) |
|  | Financial Burden Beyond Insurance | 122 (4.7%) |
|  | Care Coordination Challenges (referrals, records) | 205 (7.9%) |
|  | Clinical Care Errors / Safety Events | 46 (1.8%) |
| Medication Experiences & Side Effects | Medication Effectiveness / Response | 656 (25.2%) |
|  | Medication Side Effects / Adverse Events | 527 (20.2%) |
|  | Infusion Experience (e.g., infusion centers, scheduling) | 82 (3.2%) |
|  | Treatment Recommendations | 435 (16.7%) |
|  | Supplements Or Alternative Therapies | 152 (5.8%) |
|  | Treatment Adherence Challenges (forgetting, intolerance) | 161 (6.2%) |
|  | Biologics vs Traditional Medications | 120 (4.6%) |
|  | Clinical Trials Or Research Participation | 23 (0.9%) |
|  | Other | 30 (1.2%) |
| Symptoms & Physical Experiences | Acute or Worsening Flares | 785 (30.2%) |
|  | Fatigue | 642 (24.7%) |
|  | Pain | 969 (37.2%) |
|  | Skin Manifestations (rashes, photosensitivity) | 632 (24.3%) |
|  | Cognitive Symptoms (brain fog, memory) | 183 (7.0%) |
|  | Other Symptom Presentation (e.g., renal, cardio) | 980 (37.6%) |
|  | Other | 82 (3.2%) |
| Lifestyle & Self-Management | Symptom Management Strategies | 569 (21.9%) |
|  | Diet and Nutrition Changes | 190 (7.3%) |
|  | Exercise Adaptations | 111 (4.3%) |
|  | Stress Management Strategies | 124 (4.8%) |
|  | Workplace Disclosure / Accommodations & Disability Benefits | 210 (8.1%) |
|  | Sun Exposure Management | 133 (5.1%) |
|  | Self-Tracking / Tools (journals, apps) | 71 (2.7%) |
|  | Activities Of Daily Living Adjustments | 479 (18.4%) |
|  | Body Image / Identity Adjustments | 162 (6.2%) |
|  | Reproductive / Fertility Treatment Considerations | 98 (3.8%) |

*GPT, Generative Pre-trained Transformer*

## RESULTS

### Collection of SLE related posts

Among the 4633 collected posts, 2,030 (43.8%) were excluded according to the predefined data preprocessing pipeline (Figure 1). Exclusions included posts with missing author identifiers or insufficient title clarity (n = 252), duplicate entries (n = 1,013), and promotional and other non-patient reported content (n = 765) (Figure 1). The final dataset consisted of 2,603 unique posts (56.2%), of which 2,564 (98.4%) published after 2020 (Table 3).

**Table 3.** Number of posts by publication year.

| Year | Number of Posts |
| --- | --- |
| 2011 | 8 |
| 2012 | 3 |
| 2013 | 9 |
| 2014 | 1 |
| 2015 | 2 |
| 2016 | 2 |
| 2017 | 3 |
| 2018 | 5 |
| 2019 | 8 |
| 2020 | 33 |
| 2021 | 33 |
| 2022 | 46 |
| 2023 | 71 |
| 2024 | 278 |
| 2025 | 2,101 |

### LLMs consistency and performance

Inter-model agreement achieved high consistency across main themes and subthemes (Table 1 and Figure 2). Overall, percent agreement and Cohen’s κ indicated substantial agreement between Gemini 3.0 and GPT-5.2 for main themes (percent agreement = 91.04%; mean κ = 0.71; range = 0.42 – 0.88) and for subthemes (percent agreement = 94.43%; mean κ = 0.71; range = 0.16 – 0.91).

At the main-theme level, almost perfect agreement was observed for *Medication Experiences & Side Effects* (percent agreement = 94.08%; κ = 0.88). Substantial agreement was also observed for *Healthcare System Barriers* (percent agreement = 89.01%; κ = 0.76), *Lifestyle & Self-Management* (percent agreement = 85.79%; κ = 0.71), *Diagnosis Journey & Uncertainty* (percent agreement = 94.08%; κ = 0.74), and *Symptoms & Physical Experiences* (percent agreement = 92.24%; κ = 0.76). In contrast, *Emotional & Social Support* demonstrated moderate agreement despite high percent agreement (percent agreement = 98.27%; κ = 0.42).

Across the 43 subthemes, 15 subthemes demonstrate almost perfect agreement (κ range = 0.81 – 0.91). The highest agreement was observed for *Insurance Challenges* (e.g. *Coverage, Prior Auth*o*rization*), *Fatigue, and Cognitive Symptoms* (e.g. *Brain Fog, Memory*), each achieving κ = 0.91 with percent agreement of 99.35%, 96.77%, and 98.77%, respectively. Among the remaining subthemes, 17 demonstrated substantial agreement, four demonstrated moderate agreement, one demonstrated fair agreement and two demonstrated slight agreement (Table 1).

Post-level agreement analyses further supported high inter-model consistency. Across the six main themes, 75% of posts exhibited greater than 80% agreement between the two LLMs (Figure 2a). *Healthcare System Barriers* showed the highest consistency, with more than 75% of classified posts achieving 100% agreement. For the remaining main themes, more than half of posts demonstrated 100% agreement at the subtheme level (Figure 2b).

Compared with the consensus reference labels derived from 100 randomly selected posts, GPT-5.2 achieved higher overall performance than Gemini 3.0 (F1: 0.844 vs 0.811), driven by higher precision (0.743 vs 0.694) with similarly high recall (0.977 vs 0.975). Across the 36 subthemes that have sampled posts in our human evaluation, GPT-5.2 outperformed Gemini 3.0 in ten subthemes, Gemini 3.0 outperformed GPT-5.2 in five subthemes, and the models performed comparably in 21 subthemes. Both models demonstrated very high recall, indicating strong sensitivity for detecting relevant main themes and subthemes, while GPT 5.2 showed higher precision, resulting in a higher F1 score and a better balance between sensitivity and false-positive control. We therefore chose GPT-5.2 as the best-performing model and conducted the following thematic analysis using it.

### Thematic distribution from the best-performing model

Table 3 summarizes the distribution of main themes and subthemes identified by GPT-5.2. Of the 2,603 analyzed posts, each post received at least one subtheme label, with a mean of approximately six subthemes assigned per post.

The most frequent subtheme was *Advice Seeking* (84.1%), followed by *Emotional Coping* (e.g., *venting, hope, resilience*) (55.6%) within *Emotional & Social Support*. Discussions within *Symptoms & Physical Experiences* were also common, including *Other Symptom Presentation* (e.g., *renal, cardio*, 37.6%), *Pain* (37.2%), and *Acute or Worsening Flares* (30.2%). Within the *Diagnosis Journey & Uncertainty*, *Emotional Impact of Uncertainty and Disease Progression* was frequently labeled (33.0%). Treatment-related content was prevalent as well, notably *Medication Effectiveness/Response* (25.2%).

### Qualitative review of patient narratives

To contextualize the quantitative distribution, we conducted a qualitative review of representative patient narratives (Figure 3). Several narratives described difficulty navigating referrals, records, and care coordination. For example, one patient shared an uncertain diagnosis (“*it could be lupus*…without no follow-up plan”), and another portrayed the path to diagnosis as a prolonged “*puzzle*,” as the journey spanning multiple years (“*battling for 6 years just to be diagnosed.*”).

Many posts reflected reliance on peer communities for validation and guidance, particularly after perceived dismissal in clinical encounters or during delays in accessing specialty care. For example, one patient wrote, “*Please let me know y’alls opinions”* regarding whether beta-blocker use could be related to drug-induced lupus. Another solicited community opinions on interpreting laboratory results (“*low, total immunoglobulin E… what it means in relation to what I have going on?*”) while a long awaiting clinical appointment (“*Don’t see the doctor for another 2 months*”).

Beyond informational needs, numerous posts captured emotional distress and identity challenges, including self-doubt and feelings of invalidation (e.g., “*I’m not sure what’s wrong with me*”), as well as guilt related to role functioning when symptoms interfered with caregiving or work responsibilities (“*Feel like a failure of a mother…can’t even hold myself up most days.*”).

Narratives emphasizing multisystem involvement and functional impairment were common. One patient described that lupus “*doesn’t pick one system; it attacks them all,*” alongside accounts of symptoms being minimized as “*just a rash.*” Accounts of severe, diffuse pain and heightened sensitivity appeared frequently (“*Everything hurt.*”). In discussions of acute or worsening flares, posters described abrupt escalation of symptoms and extended recovery periods, including delayed sequelae (e.g., “*hair loss lagging 2–3 months after severe systemic flare.*”).

Posts also conveyed anxiety driven by ambiguous diagnostic pathways and fragmented care planning. For instance, one patient described cross-specialty deferral during evaluation of acute foot pain, stating that the podiatrist *“refused to treat my foot pain”* and told the patient, *“I don’t treat lupus.”* Finally, posts reflected uncertainty regarding whether treatments were working and how quickly improvement should occur, illustrated by posts such as “*Monthly ER visits since June…each triggered by new infections due to immunosuppression*” and “*Torn between fear of flares if dose drops vs. fear of toxicity if it stays high*”, which demonstrated current unmet medical needs of SLE patients.

## DISCUSSION

This study applied LLMs to analyze lupus-related Reddit posts and identify patient-expressed unmet medical needs and lived experiences in SLE. Our finding indicated that patients described a multidimensional spanning from diagnosis uncertainty, emotional and social needs, healthcare system barriers, treatment experiences, symptom burden, and self-management challenges. Advice seeking and emotional coping were the most prominent narrative patterns, suggesting substantial reliance on peer networks particularly when patients feel unheard in clinical settings. These psychosocial needs align with prior qualitative studies describing ongoing gaps in communication, validation, and daily functioning among individuals living with SLE.^24,25^

Specific symptom experiences shared by patients corresponded with established qualitative literature emphasizing patient-centered voices on pain^26^, fatigue^27,28^, acute or worsening flares^29^. Cognitive symptoms, including brain fog and memory difficulties, were also commonly discussed, consistent with previous qualitative research on lupus-related cognitive impairment.^30^ Notably, Other symptom presentations was one of the most prevalent subthemes, reinforcing that SLE rarely exists in isolation with its impact often amplified by coexisting chronic conditions. These observations were supported by a prior study focused on describing the impact of comorbidities on quality of life in SLE^31^ and underscore the need to approach disease burden as prioritized by patients, rather than focusing on a narrow set of symptoms.

Treatment experiences emphasized unmet medical needs in medication decision-making and long-term treatment management, including uncertainty about effectiveness, concerns about adverse effects, and adherence challenges. Prior studies have described medication concerns and adherence challenges driven by the fear of side effects, forgetting, disbelief in medication,^32^ while identifying trust in rheumatologists and structured follow-up as key facilitators of sustained adherence.^33^ Similarly, patient narratives in our results reflected uncertainty regarding medication effectiveness and drug-drug interactions, as well as barriers to adherence, suggesting that SLE management often involves medication regimens that may seem as complex as the disease itself with limited clarity on long-term impacts.

Moreover, unmet medical needs related to uncertainty and concerns about disease progression were also frequently observed within the diagnosis journey, consistent with prior findings that uncertainty may persist even after diagnosis.^34^ Provider trust and distrust emerged as an important contextual factor for previously described medication adherence facilitators^33^, as early clinical interactions may shape patients confidence in treatment recommendations and engagement with longitudinal care. Delayed diagnosis or misdiagnosis were commonly reported, often in the context of symptom minimization or misattribution, aligning with qualitative studies across diverse geographic settings that have examined barriers and facilitators associated with timely diagnosis and appropriate treatment in SLE.^35–38^

While diagnostic uncertainty^34^ and challenges in patient–provider dynamics^32,33,39^ have been well documented in previous qualitative studies of SLE patient experience, confusion related to serologic testing of patient experience during the diagnostic process has received comparatively limited attention in prior qualitative and social media-based studies. In this study, patient confusion surrounding interpretation of laboratory test results, such as anti-nuclear antibody (ANA) and complement levels, appeared as a frequent unmet medical need. This knowledge gap often drove patients to seek peer interpretations of laboratory values and discordance between symptoms and test results.

Taken together, our findings were largely consistent with existing literature from multiple main themes on unmet medical needs and patient-reported experience in SLE while also highlighting persistent gaps in patient education and communication between clinician and patients that warrant greater attention in patient-centered care. The consistency of major unmet medical need themes with prior literature also supports the credibility of LLM based thematic analysis as a complementary approach for synthesizing large scale unstructured patient narratives from social media.

This study had several limitations. First, the analysis is subject to self-selection bias inherent to social media research. Users who post in lupus-related subreddits may not represent the broader SLE population and may have higher symptom burden, greater unmet needs, or stronger engagement with online peer communities than those who do not participate in online forums.^40^ Consequently, the narratives captured may overrepresent more persistent, disruptive, or emotionally salient experiences. Second, the anonymity of the Reddit platform precluded the verification of users’ clinical diagnoses and restricted access to sociodemographic information.^41^ As a result, our analysis relied on self-identified SLE status, which could not be independently confirmed and may have included individuals with other lupus-related disorders. This limitation is inherent to social media research and should be considered when interpreting the generalizability of our findings. Furthermore, subgroup analyses by age, sex, race or ethnicity, or geographic were not feasible, and variation in unmet needs across social determinants of health could not be assessed.

Third, the analysis was restricted to a single social media platform. Although additional lupus-related platforms were explored, data access restrictions and usage policies limited compliant large-scale extraction. Finally, while LLMs offer scalable analysis of unstructured data, their outputs may vary across repeated runs.^42^ We observed approximately 10% variation in classification during automated identification of promotional and non-patient reported posts, consistent with prior evidence of stochastic variability in LLM outputs, analogous to inter-rater variability in human annotation.

## CONCLUSION

Our study demonstrates that large-scale social media listening, enabled by state-of-the-art LLMs, can provide timely, patient-centered insights into unmet medical needs and lived experiences in SLE. Consistent with prior qualitative and survey-based studies, patient narratives reflected a multidimensional burden spanning diagnostic uncertainty, symptom and treatment challenges, emotional distress, and gaps in clinical support and communication. By systematically analyzing thousands of patient-generated narratives, this work illustrates an efficient and scalable approach to synthesizing unstructured data that complements established patient-focused methods. More broadly, the analytical framework presented here can be extended to other disease areas to support rapid identification of patient priorities, inform patient-centered care strategies, and guide evidence generation relevant to therapeutic development and health services research.

## Contributions

SY and LY conceived and designed the study and drafted the manuscript. LY supervised the study. CH, NE, and JO conducted qualitative review and manual annotations. JL conducted data collection, preprocessing, and LLM-based thematic classification. SY conducted the data analysis. ER provided clinical review and critical revisions. All authors reviewed and approved the final version.

## Data availability statement

The data analyzed in this study were derived from publicly available Reddit posts accessed via the Reddit API. Processed datasets and analysis codes may be made available by the corresponding author upon reasonable request, subject to platform terms and privacy considerations.

